# Chromatin remodeler *developmental pluripotency associated factor 4* (*DPPA4*) is a candidate gene for alcohol-induced developmental disorders

**DOI:** 10.1101/2022.04.14.22273502

**Authors:** P Auvinen, J Vehviläinen, H Marjonen, V Modhukur, J Sokka, E Wallén, K Rämö, L Ahola, A Salumets, T Otonkoski, H Skottman, M Ollikainen, R Trokovic, H Kahila, N Kaminen-Ahola

## Abstract

Alcohol affects embryonic development, causing a variable fetal alcohol spectrum disorder (FASD) phenotype with neuronal disorders and birth defects. To explore the etiology of FASD, we collected an exceptional cohort of 80 severely alcohol-exposed and 100 control newborns and performed genome-wide DNA methylation and gene expression analyses of placentas. *DPPA4, FOXP2*, and *TACR3* with significantly decreased DNA methylation were discovered – particularly the regulatory region of *DPPA4* in the early alcohol-exposed placentas. When human embryonic stem cells (hESCs) were exposed to alcohol *in vitro*, significantly altered regulation of *DPPA2*, a closely linked heterodimer of *DPPA4*, was observed. While the regulatory region of *DPPA4* was unmethylated in both control and alcohol-exposed hESCs, alcohol-induced decreased DNA methylation similar to placenta was seen in *in vitro* differentiated mesodermal and ectodermal cells. Furthermore, common genes with alcohol-associated DNA methylation changes in placenta and hESCs were linked exclusively to the neurodevelopmental pathways, which emphasizes the value of placental tissue when analyzing the effects of prenatal environment on human development. Our study shows the effects of early alcohol exposure on human embryonic and extraembryonic cells, introduces candidate genes for alcohol-induced developmental disorders, and reveals potential biomarkers for prenatal alcohol exposure.

## BACKGROUND

Prenatal alcohol exposure (PAE) is associated with a broad spectrum of permanent structural, physiological, neurocognitive, and behavioral disorders of the exposed, often growth restricted offspring^1^. Fetal alcohol spectrum disorders (FASD) are a consequence of PAE, and an umbrella term for all alcohol-induced developmental disorders. PAE is one of the most harmful environmental factors affecting permanently 3–5% of individuals in the Western world^2^.

Several lines of evidence suggest that the epigenome of developing embryo is sensitive to environmental effects in the beginning of pregnancy, during the dynamic period of epigenetic reprogramming^3,4^. Alcohol-induced epigenetic alterations have been observed in the offspring of our early PAE mouse model^5,6^ as well as human and mouse embryonic stem cells^7,8^. Those early epigenetic changes could affect gene regulation and consequently developmental programming. Depending on the function of the cell or tissue types, they could contribute to the complex phenotype of FASD.

To explore the etiology of FASD we have collected placental samples from PAE and control newborns at birth. Placenta is an accessible human tissue and a promising implement for identifying the effects of intra-uterine environment on embryonic development, including neuronal development^9,10^. Here, by performing genome-wide DNA methylation (DNAm) analysis of placenta, we discovered a candidate gene *developmental pluripotency associated factor 4* (*DPPA4*), which was hypomethylated particularly in the early alcohol-exposed placentas. *DPPA4* functions as a heterodimer with *developmental pluripotency associated factor 2* (*DPPA2*) and both proteins are required for efficient binding and chromatin remodeling^11^. By modifying chromatin structure, these epigenetic priming factors facilitate transition between pluripotency and differentiation^11,12,13^, which makes them plausible candidate genes for developmental disorders. Both genes are located in tandem on chromosome 3q13.13, they are regulated by promoter DNAm in mouse, and are expressed for a short time in the beginning of embryonic development^14^. To explore the effects of early alcohol exposure on human embryonic cells, and more specifically on the regulation of *DPPA2* and *DPPA4*, we performed genome-wide DNAm and gene expression analyses for *in vitro* alcohol-exposed human embryonic stem cells (hESCs). Furthermore, hESCs were *in vitro* alcohol-exposed during differentiation into the endodermal, mesodermal, and ectodermal cells.

Alcohol-induced epigenetic changes in the first embryonic cells could be fixed in persistent cellular memory and mitotically transmitted to different cell and tissue types. Indeed, variety of PAE-associated DNAm changes in peripheral blood^15^ and buccal epithelial cells (BECs)^16,17^ of children with FASD have been observed in previous genome-wide studies. Therefore, we examined whether PAE-associated epigenetic alterations can be detected not only in the extraembryonic placenta, but also in embryonic white blood cells (WBCs) from cord blood or BECs of the same newborns. Those changes could be the first fingerprints of PAE, potential future biomarkers for FASD, which would enable early diagnosis and personalized support for the development of the affected children.

## RESULTS

### Characteristics of epigenetics of FASD (epiFASD) cohort

General characteristics of epiFASD cohort including 80 PAE and 100 control newborns as well as their mothers were compared (Table 1, Supplementary Tables 1 and 2). The newborns’ birth weight, length and head circumference (HC) were compared using standard deviations (SDs) of measures based on international growth standards, in which the gestational age at birth and sex are considered^18^. PAE newborns had significantly smaller birth weights (SD), lengths (SD) and HCs (SD) compared to control newborns (*P* = 0.030, Mann-Whitney U, *P* = 0.044 and *P* = 0.012, Student’s *t*-test, respectively). Moreover, to explore the effects of early PAE, we selected 28 newborns whose mothers had consumed alcohol up to gestational week seven at maximum to the separate analyses. Notably, also the newborns in this early PAE subgroup had significantly smaller HCs (SD) compared to controls (*P* = 0.018, Student’s *t*-test).

**Table 1.** General characteristics of PAE and control newborns as well as their mothers included in the phenotype analysis. Differences in weight, length, and head circumference (HC) of the newborns were calculated using both anthropometric measures and the SDs of measures based on international growth standards adjusted for gestational age at birth and gender. Data presented as mean ±SD. *P*-values for PAE and control newborns as well as for early PAE and control newborns were calculated using Mann-Whitney U test (1) or two-tailed Student’s *t*-test (2) and for the proportion of males and females between the groups using Pearson chi-square (3). Differences in gestational age were calculated for all newborns and for newborns delivered spontaneously without cesarean section (CS) and induction of labor (IL). ILs due to prolonged control pregnancies (42+1) were not excluded.

| Newborns | Sex<br>(male/<br>female) | Gestational<br>age<br>(weeks +days) |  | Birth weight |  | Birth length |  | HC |  | Placental<br>weight |
| --- | --- | --- | --- | --- | --- | --- | --- | --- | --- | --- |
|  |  | all | without<br>CS/IL | (g) | SD | (cm) | SD | (cm) | SD | (g) |
| **Control (n = 100)**<br>**mean (±SD)** | 52.0/<br>48.0% | 40 +3<br>(±1 +0) | 40 +4<br>(±1 +0)<br>n = 90 | 3661<br>(±366) | 0.07<br>(±0.9) | 51<br>(±2) | -0.24<br>(±0.7) | 35.5<br>(±1.3) | 0.25<br>(±0.9) | 626.5<br>(±126.4) |
| **PAE (n = 80)**<br>**mean (±SD)** | 42.5/<br>57.5% | 39 +4<br>(±1 +5) | 39 +4<br>(±1 +6)<br>n = 60 | 3327<br>(±554) | -0.23<br>(±0.1) | 49<br>(±3) | -0.50<br>(±0.8) | 34.4<br>(±1.9) | -0.14<br>(±1.2) | 596.6<br>(±136.7) |
| **Difference between**<br>**groups, P-value** | 0.231<sup>(3)</sup> | <.001<sup>(1)</sup> | <.001<sup>(1)</sup> | <.001<sup>(1)</sup> | 0.030<sup>(1)</sup> | <.000<sup>(1)</sup> | 0.044<sup>(2)</sup> | <.000<sup>(1)</sup> | 0.012<sup>(2)</sup> | 0.083<sup>(1)</sup> |
| **Early PAE (n = 28)**<br>**mean (±SD)** | 46.4/<br>53.6% | 40 +1<br>(±1 +4) | 40 +2<br>(±1 +3)<br>n = 24 | 3523<br>(±433) | -0.05<br>(±0.8) | 50<br>(±2) | -0.41<br>(±0.8) | 34.5<br>(±1.7) | -0.23<br>(±1.2) | 616.9<br>(±158.4) |
| **Difference between**<br>**groups, P-value** | 0.672<sup>(3)</sup> | 0.659<sup>(1)</sup> | 0.717<sup>(1)</sup> | 0.131<sup>(2)</sup> | 0.489<sup>(2)</sup> | 0.233<sup>(2)</sup> | 0.344<sup>(2)</sup> | <.001<sup>(1)</sup> | 0.018<sup>(2)</sup> | 0.421<sup>(1)</sup> |

| Mothers | Age | Parity | Pre-pregnancy<br>BMI | Gestatio-<br>nal weight<br>gain |
| --- | --- | --- | --- | --- |
|  | (years) |  | (kg/m2) | (kg) |
| **Control (n = 100)**<br>**mean (±SD)** | 31.6<br>(±4.6) | 0.55<br>(±0.7) | 22.8<br>(±3.5)<br>n = 99 | 15.1<br>(±3.8)<br>n = 88 |
| **PAE (n = 80)**<br>**mean (±SD)** | 29.5<br>(±6.8) | 0.66<br>(±1.3) | 25.4<br>(±5.9)<br>n = 67 | 12.4<br>(±6.7)<br>n = 56 |
| **Difference between**<br>**groups, P-value** | 0.017<sup>(1)</sup> | 0.269<sup>(1)</sup> | 0.008<sup>(1)</sup> | 0.008<sup>(2)</sup> |
| **Early PAE (n = 28)**<br>**mean (±SD)** | 27.4<br>(±6.0) | 0.36<br>(±0.8) | 24.1<br>(±4.9)<br>n = 23 | 15.4<br>(±6.9)<br>n = 17 |
| **Difference between**<br>**groups, P-value** | 0.001<sup>(2)</sup> | 0.046<sup>(1)</sup> | 0.313<sup>(1)</sup> | 0.215<sup>(2)</sup> |

When potential correlations between changes in placental weight (g), birth measures (SDs), and maternal alcohol consumption determined by AUDIT scores^19^ or the number of alcohol units per week were calculated between the PAE as well as early PAE newborns and controls, a negative correlations between AUDIT scores and birth length (SD) were found (*r* = -0.505, *P* < 0.001, *n* = 44, *r* = -0.576, *P* = 0.012, *n* = 18, Spearman’s rank correlation, respectively). The gestational age was significantly shorter in PAE pregnancies compared to the controls (*P* < 0.001, Mann-Whitney U). Furthermore, the mothers of PAE newborns had significantly higher pre-pregnancy BMI, but gained significantly less weight during pregnancy than the mothers of control newborns (*P* = 0.008, Student’s t-test and *P* = 0.008, Mann-Whitney U, respectively). However, the weight gain in both groups was within the recommended range^20^.

### Effects of PAE on genome-wide DNAm in the placenta

We used Illumina’s Infinium MethylationEPIC microarrays to identify PAE-associated genome-wide DNAm alterations in 69 PAE and 66 control full-term placentas (general characteristics in Supplementary Table 3). By adjusting for batch, sex, and smoking covariates the analysis resulted in 2,538 significantly differentially methylated CpG sites (2,138 hypomethylated and 400 hypermethylated) with false discovery rate (FDR) < 0.05 (Fig. 1a,b and Supplementary Table 4). To separate the most prominent changes, we focused on the CpG sites with DNAm difference of > 5% between PAE and control placentas, which we considered as biologically significant. There were 689 biologically significant CpG sites (hereafter differentially methylated positions, DMPs) associated with PAE (FDR < 0.05, -0.05 > Δ *β* > 0.05), of which 481 were hypomethylated and 208 hypermethylated.

**Fig. 1.**
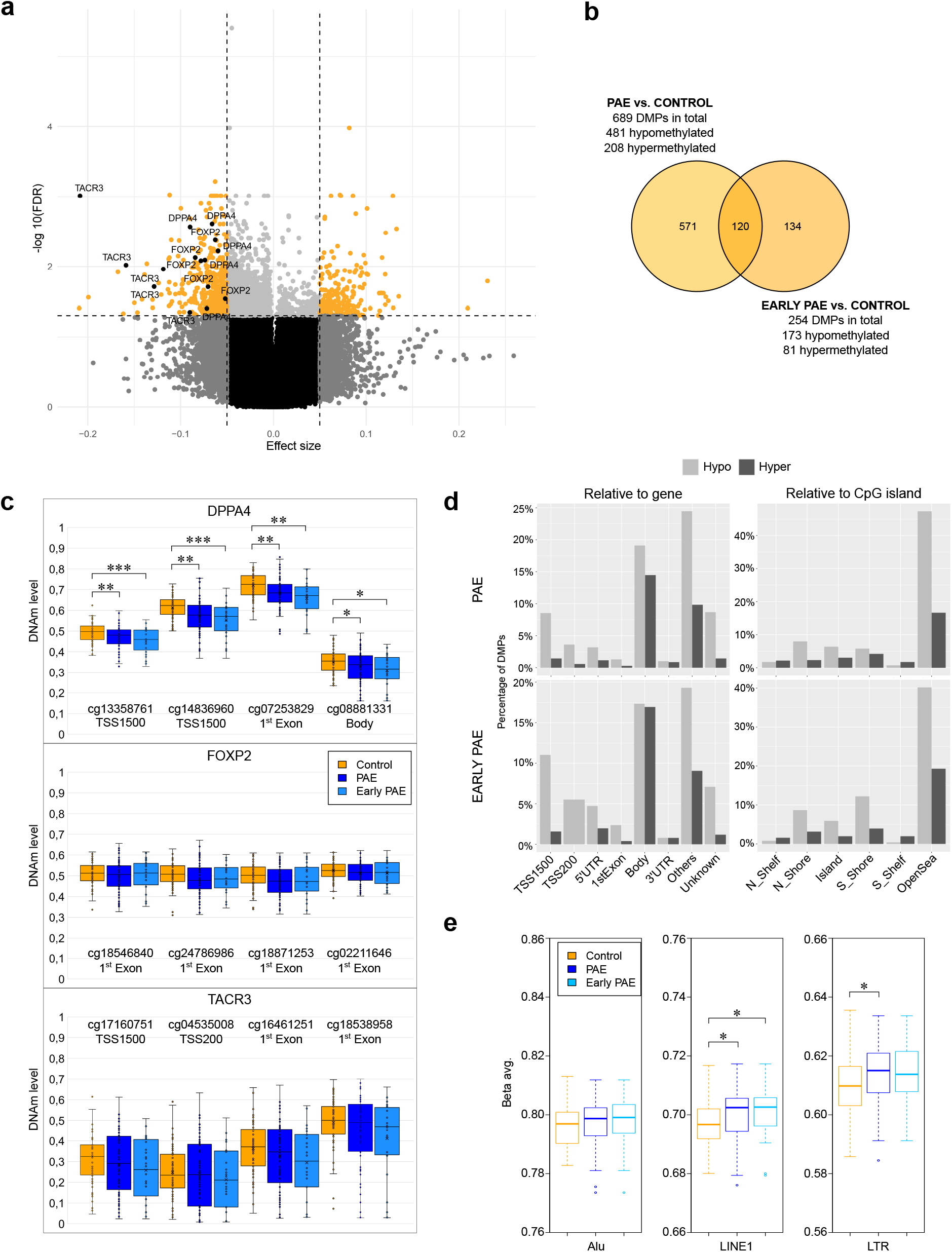
PAE-associated differential DNAm in the placenta. **a** Volcano plot showing the distribution of associations between placental CpG sites and PAE. Horizontal line marks FDR 0.05 and vertical line marks effect size ± 0.05. **b** Venn diagram showing the number of PAE-associated DMPs, which are in common between all PAE placentas and the early PAE subgroup. **c** DNAm levels of DMPs with the largest Δβ-values and their locations in relation to gene in *DPPA4, FOXP2*, and *TACR3* in control, PAE, and early PAE placentas. \**P* < 0.05, \*\**P* < 0.01, \*\*\**P* ≤ 0.001, two-tailed Student’s *t*-test. **d** Genomic location of PAE- and early PAE-associated DMPs in relation to gene and CpG island in the placenta. DMPs were divided to hypo- and hypermethylated subgroups, which were further grouped according to the genomic location according to UCSC database. If the location information was missing, DMP was marked as ‘unknown’. In case of multiple location entries, group ‘others’ was used. **e** Effects of PAE on global placental DNAm level predicted by Alu, LINE1, and LTR repetitive regions in all PAE placentas and in the early PAE subgroup. \**P* < 0.05, two-tailed Student’s *t*-test. Placental samples: control *n* = 66, PAE *n* = 69, and early PAE *n* = 28. Abbreviations TSS1500: 1500 bp upstream of transcription start site, TSS200: 200 bp upstream of TSS, UTR: untranslated region, N_shelf: north shelf, N_shore: north shore, S_shore: south shore, S_shelf: south shelf.

The analysis revealed five hypomethylated DMPs in *DPPA4*. Noteworthily, the DNAm level of the most significantly altered DMP (cg14836960) was below 0.5 (50%) in 10 PAE placentas, whereas DNAm level was 0.5 or above in all control placentas (Fig. 1c). Furthermore, *transcription factor forkhead box P2* (*FOXP2*), which is needed for the development of speech regions in the brain during embryogenesis^21,22^ had six hypomethylated DMPs, and *Tachykinin Receptor 3* (*TACR3 or neurokinin 3 receptor, NK3R*) expressed in the central nervous system, had five hypomethylated DMPs. Observed DMPs were mainly located in regulatory regions and in the first exons (Fig. 1c and Supplementary Table 4). Interestingly, genetic polymorphisms in *TACR3* have been previously associated with alcohol and cocaine dependence^23^. In addition to testing for associations for each CpG separately, we tested for differentially methylated regions (DMRs) defined as a region with maximal allowed genomic distance of 1,000 bp containing three or more CpGs of which at least one CpG with a Δβ ≥ 0.05. A total of 112 DMRs were observed, including highly significant DMRs in *DPPA4* (6 CpGs), *FOXP2* (7 CpGs), and *TACR3* (10 CpGs) (Supplementary Table 5). Regarding the retarded growth associated with FASD as well as our previous finding on the genotype-specific effects of PAE on *IGF2/H19* locus DNAm in the placenta^24^, also the hypomethylated DMR in *IGF2/IGF2AS* (21 CpGs) observed in the current genome-wide study with a larger sample size is highly interesting.

We also performed genome-wide DNAm analyses of placentas in the early PAE subgroup (*n* = 28). The analysis revealed 254 PAE-associated DMPs (173 hypomethylated and 81 hypermethylated, FDR < 0.05, -0.05 > Δ*β* > 0.05) (Fig. 1b and Supplementary Table 6), including hypomethylated DMPs in *DPPA4* and *TACR3* (two and six DMPs, respectively). Noteworthily, there was a significant difference between control and the early PAE subgroup in the group mean DNAm levels of *DPPA4*, which indicates the prominent effect of early exposure (cg13358761: *P* < 0.001, cg14836960: *P* = 0.002, and cg07253829: *P* < 0.001, cg08881331: *P* < 0.019, respectively, Student’s *t*-test) (Fig. 1c). Indeed, in the group of 10 PAE placentas with DNAm level below 0.5, seven were included in the early PAE subgroup. DMR analysis for the early PAE subgroup revealed 29 DMRs, including *DPPA4* (4 CpGs) and *TACR3* (10 CpGs) (Supplementary Table 7).

Prominent PAE-associated hypomethylation of DMPs was observed in the majority of genomic locations relative to gene and CpG island in all PAE placentas – especially in the regulatory regions of placentas in the early PAE subgroup (Fig. 1d). The effect of PAE on global placental DNAm level was predicted by comparing the mean DNAm level of CpGs in Alu, LINE1, and LTR repetitive element regions, which comprise 36% of human genome in total^25,26^. On contrary to the hypomethylation observed when using all DMPs, significant hypermethylation was observed at LINE1 and LTR regions in all PAE placentas (*P* = 0.019 and *P* = 0.02, respectively, Student’s *t*-test) and in LINE1s in the early PAE subgroup (*P* = 0.029, Student’s *t*-test) compared with controls (Fig. 1e).

Pathway analyses were performed to get a comprehensive picture on the biological processes in which PAE-associated DMPs cluster. Gene Ontology (GO) enrichment analysis of PAE-associated DMPs revealed interesting biological process (BP) involved in the function of heart and nervous system as well as adult behavior (*P* < 0.05) (Fig. 2a and Supplementary Table 8). DMRs cluster to various biological processes, such as the regulation of chemotaxis, embryonic placenta morphogenesis, Wnt signaling in stem cell proliferation as well as putamen and caudate nucleus development (Fig. 2b and Supplementary Table 9). Both DMPs and DMRs cluster to the GO terms involved in vocalization behavior and genomic imprinting.

**Fig 2.**
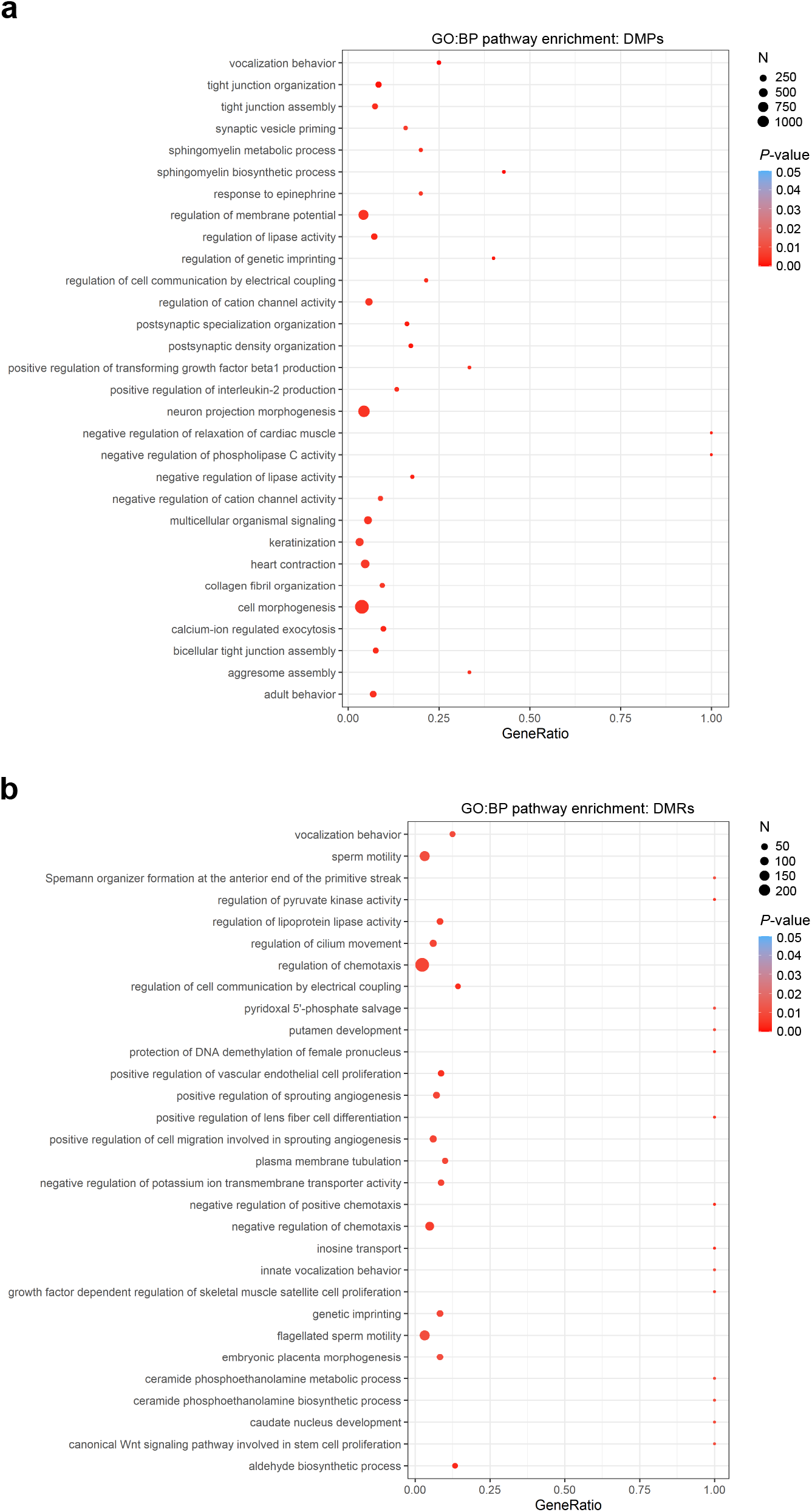
Pathway analyses of placental DMPs and DMRs. Significantly enriched terms identified in GO:BP enrichment analysis of PAE-associated **a** DMPs and **b** DMRs in placenta (*P* < 0.05). In both figures the 30 most significant pathways are shown.

### Potential biomarkers for PAE

To validate the results of genome-wide DNAm microarray analysis and to determine potential biomarkers for PAE, we examined DNAm profiles of *DPPA4, FOXP2*, and *TACR3* in placenta, WBCs, and BECs from each newborn by targeted methylation analysis using EpiTYPER (Agena Bioscience, Inc.). Targets for the EpiTYPER analysis were selected based on the microarrays and the significance of the identified DMPs when all PAE placentas were compared to controls. The placental samples with the largest DNAm difference between PAE and control groups were chosen for the further analysis (*DPPA4* (cg14836960) and *TACR3* (cg16461251) *P* < 0.001, *FOXP2* (with two DMPs in the same unit: cg18871253 and cg24786986) *P* = 0.002, Mann-Whitney U, Supplementary Table 10). The DNAm levels of selected DMPs in placenta measured by the microarrays vs EpiTYPER correlated significantly, and thus validated the DNAm results as well as the feasibility of the selected EpiTYPER primers (*DPPA4*: *r* = 0.856, *P* = 3.29×10^−9^, *n* = 29; *FOXP2*: *r* = 0.962, *P* = 4.72×10^−11^, *n* = 19; and *TACR3*: *r* = 0.903, *P* = 4.06×10^−6^, *n* = 15, Spearman’s rank correlation, respectively).

We next examined the applicability of observed PAE-associated placental DNAm differences for potential PAE biomarkers in blood or BECs, which are more easily accessible biological samples than placenta. The stability of DNAm levels across placenta, WBCs and BECs from each of the selected newborns was tested, but significant correlations between tissues or cell types were not observed. However, when only the PAE newborns with placental DNAm level of *DPPA4* DMP (cg14836960) below 0.5 (the observed threshold between PAE and control placentas in the microarray) were scrutinized, a trend of low DNAm level was detected also in BECs of the same newborns (Fig. 3a). Furthermore, a trend of PAE-associated hypomethylation of DMP in the first exon of *TACR3* (cg16461251) was observed also in WBCs and BECs (Fig. 3b). Notably, the DNAm difference in this specific DMP between selected control and PAE placentas was also significant in BECs analyzed by EpiTYPER (*P* = 0.009, Mann-Whitney U).

**Fig. 3.**
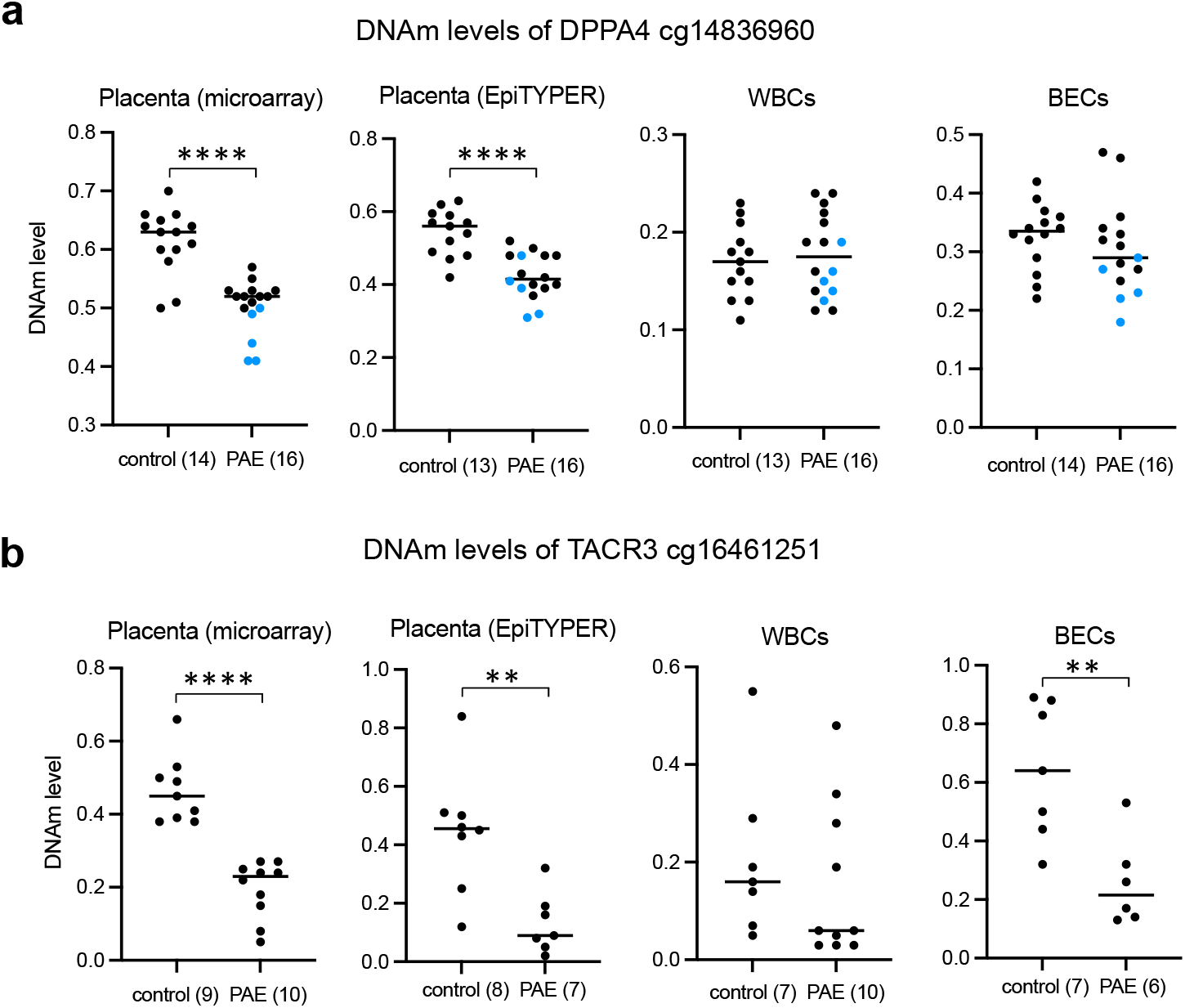
DNAm levels of *DPPA4* and *TACR3* in selected samples in the placenta, WBCs, and BECs by microarray and EpiTYPER. **a** DNAm of placental *DPPA4* DMP (cg14836960) analyzed by microarrays and EpiTYPER as well as the same CpG in WBCs and in BECs analyzed by EpiTYPER. Blue dots indicate DMP of each PAE placenta, which have DNAm level below 0.5 in microarrays. **b** DNAm of placental *TACR3* CpG (cg16461251) analyzed by microarrays and EpiTYPER as well as the same CpG in WBCs and BECs analyzed by EpiTYPER. \*\**P* < 0.01, \*\*\*\**P* < 0.0001, Mann-Whitney U.

### Effects of PAE on placental mRNA expression

To study genome-wide PAE-associated alterations in gene expression, we performed 3’mRNA sequencing (mRNA-seq) for 64 PAE and 41 control placentas (general characteristics in Supplementary Table 3). When the mRNA-seq data was adjusted by smoking and sex, we observed only one significantly differentially expressed gene, *Stress Associated Endoplasmic Reticulum Protein 1* (*SERP1*), which was upregulated in PAE placentas (FDR = 0.043). *SERP1* stabilizes membrane proteins during stress and facilitates glycosylation after stressful conditions^27^. The expressions of *IGFBP1, RBP4, DKK1, PAEP*, and *CD248* were significantly upregulated in the placentas of early PAE subgroup (*n* = 23) (Supplementary Table 11). According to our analysis, *DPPA2, DPPA4, FOXP2*, or *TACR3* are not expressed in the term placenta.

When the data was analyzed without covariate adjustments, we observed 1,076 genes with significantly altered expression in all PAE placentas (FDR < 0.05) (Fig. 4a,b and Supplementary Table 12). This may indicate a strong effect of smoking and sex on gene expression in the placenta, which mask the effects of PAE. Among the most upregulated genes were cytochrome P450 enzymes *CYP1A1* and *CYP1B1*, which are involved in xenobiotic metabolism^28^ and are also known to be induced by maternal smoking^29,30^. Also, aromatase *CYP19A1*, which is highly expressed in the placenta and catalyzes the synthesis of estrogen from androgen, is significantly upregulated in PAE placentas. The expressions of 28 genes were significantly differentially expressed in the placentas of the early PAE subgroup (FDR < 0.05) (Supplementary Table 13), and their expression was significantly altered also in all PAE placentas (Fig. 4b).

**Fig. 4.**
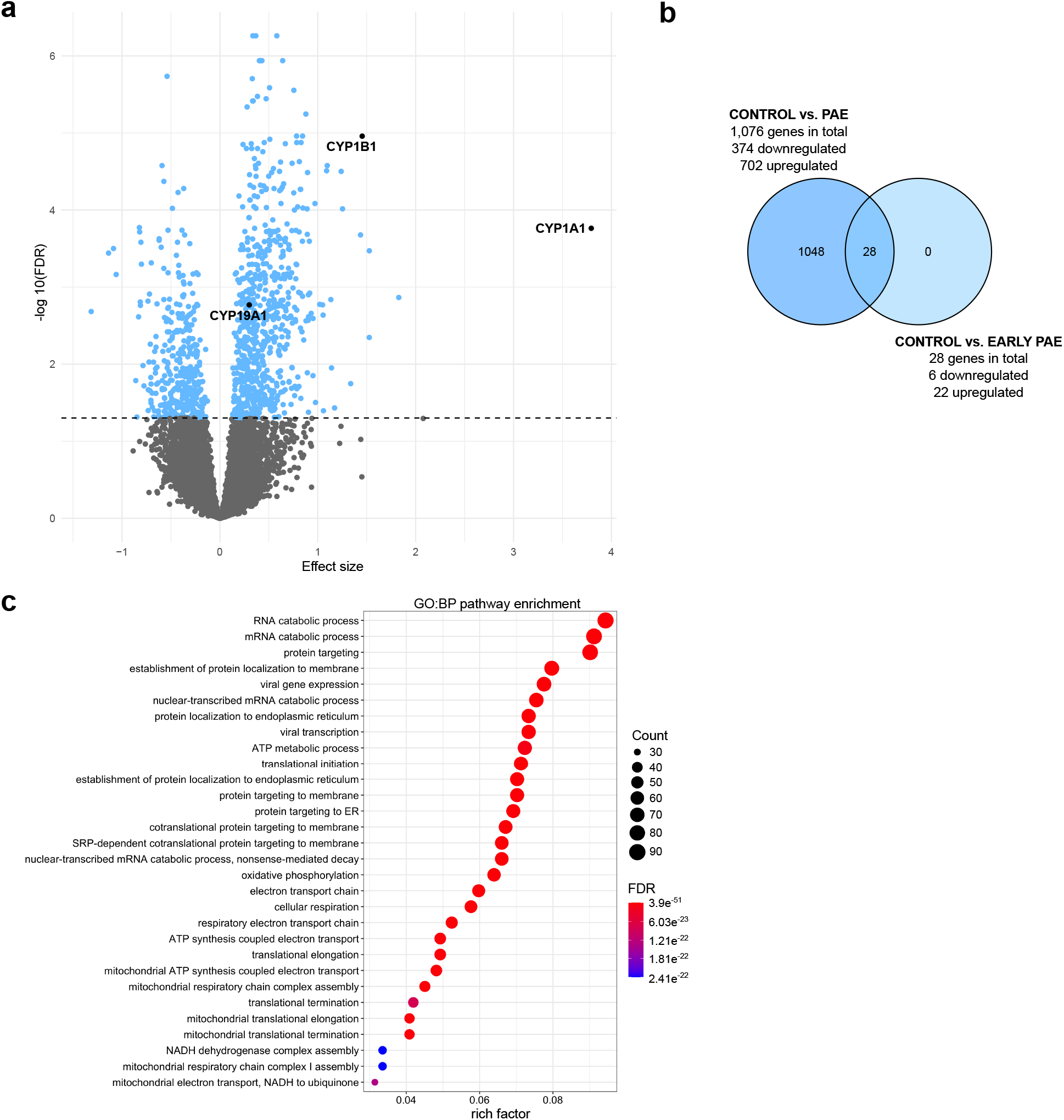
PAE-associated differential gene expression in the placenta. **a** Volcano plot showing the distribution of associations between mRNA expression and PAE. Horizontal line marks FDR 0.05. **b** Venn diagram showing the number of PAE-associated differentially expressed genes, which are in common between all PAE placentas and the early PAE subgroup. **c** Significantly enriched terms identified in GO:BP enrichment analysis of PAE-associated differentially expressed genes in placenta (FDR-corrected *q*-value < 0.05). The 30 most significant pathways are shown. Placental samples: control *n* = 41, PAE *n* = 64, and early PAE *n* = 23.

According to the GO:BP enrichment analysis, PAE-associated gene expression from the null model is linked to protein targeting to endoplasmic reticulum and cellular respiration in mitochondria (FDR-corrected *q*-value < 0.05) (Fig. 4c and Supplementary Table 14). To characterize the potential effect of PAE-associated DNAm on gene expression levels in the same samples, we performed correlation analysis. The analysis revealed eight genes, which had significant PAE-associated correlation between decreased DNAm and increased mRNA expression in the placenta: *B3GNT3, CBR1, CNDP2, HEATR5A, PRKAG2, S100A14, SAR1B*, and *TUSC3* (*P* < 0.01) (Supplementary Table 15).

### Effects of *in vitro* alcohol exposure on hESCs

To explore the effects of early alcohol exposure on hESCs without potential confounding factors associated with human studies *in vivo*, we exposed two cell lines (H1 and Regea08/017) with replicates to 70 mM alcohol for 48 hours and performed genome-wide DNAm and gene expression analyses. By using Illumina’s EPIC microarrays (eight alcohol-exposed and eight control samples) we identified 10,888 alcohol-induced differentially methylated CpG sites (10,046 hypomethylated and 842 hypermethylated) as well as 1,111 hypermethylated non-CpG sites (mCpHs) common in hESCs with FDR < 0.05 (Fig. 5a and Supplementary Table 16). Of all differentially methylated sites, 3,700 (1,879 hypomethylated CpGs, 714 hypermethylated CpGs, and 1,107 hypermethylated CpHs) were considered as biologically significant DMPs (FDR < 0.05, -0.05 > Δ*β* > 0.05) and were used in further analysis. Furthermore, a total of 442 DMRs were observed (Supplementary Table S17).

**Fig. 5.**
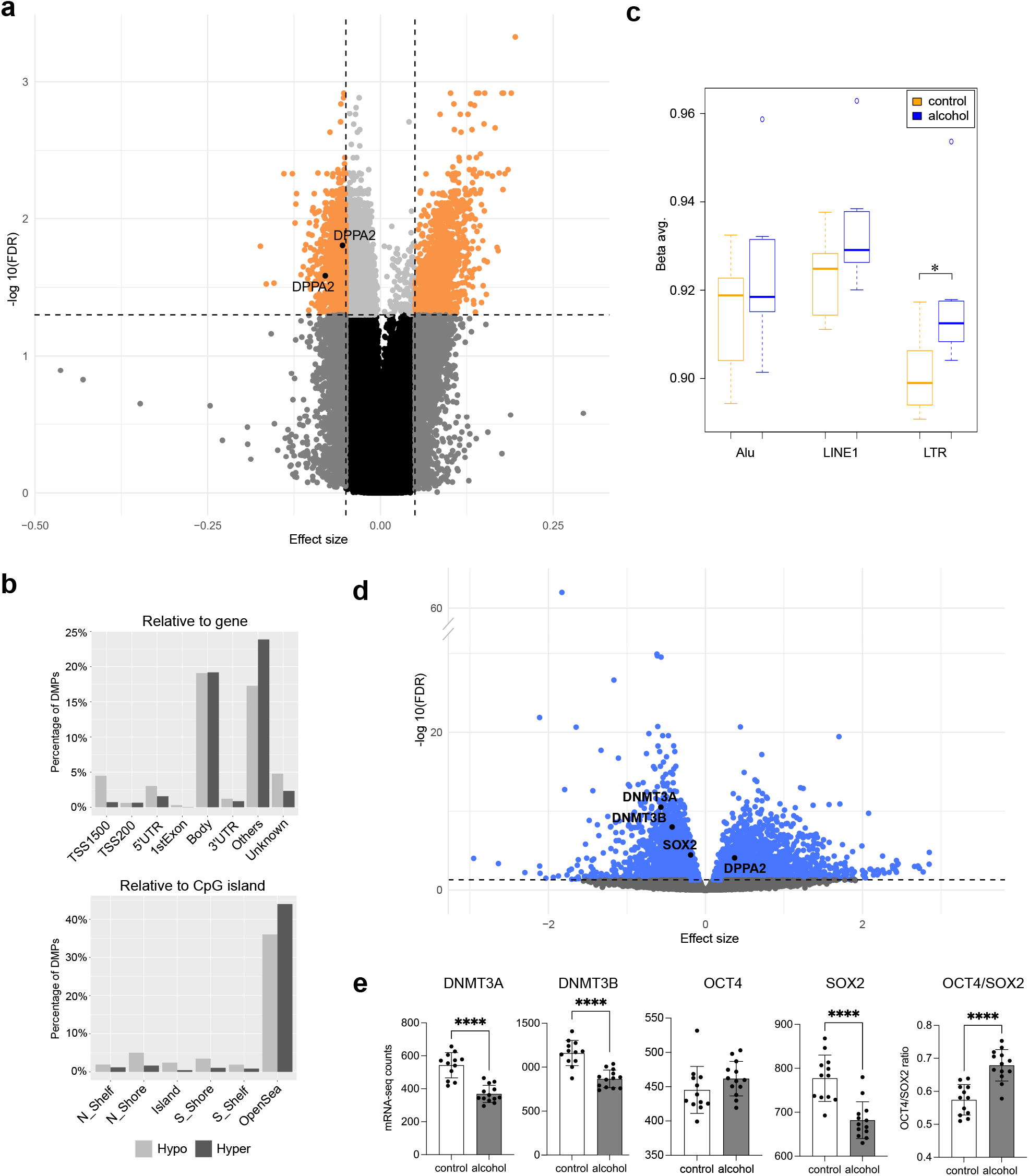
Effects of alcohol exposure on hESCs. **a** Volcano plot showing the distribution of associations between CpG sites and alcohol exposure in hESCs. Horizontal line marks FDR 0.05 and vertical line marks effect size ± 0.05. **b** Location of alcohol-induced DMPs in relation to gene and CpG island in hESCs. DMPs were divided to hypo- and hypermethylated subgroups, which were further grouped according to the genomic location according to UCSC database. If the location information was missing, DMP was marked as ‘unknown’. In case of multiple location entries, group ‘others’ was used. **c** Effects of alcohol exposure on global DNAm level in hESCs predicted by Alu, LINE1, and LTR repetitive regions. \**P* < 0.05, two-tailed Student’s *t*-test. **d** Volcano plot showing the distribution of associations between mRNA expression and alcohol exposure. Horizontal line marks FDR 0.05. **e** *DNMT3A, DNMT3B, OCT4*, and *SOX2* gene expressions as well as *OCT4*/*SOX2* expression ratio in control and alcohol-exposed hESCs. Data presented as mean ±SD. \**P* < 0.05, two-tailed Student’s *t*-test and \*\*\*\**P* ≤ 0.0001, FDR-corrected *P*-value. Control hESCs *n* = 8 and *n* = 12 as well as alcohol-exposed hESCs *n* = 8 and *n* = 13 in DNAm and mRNA-seq analyses, respectively.

The global trends of alcohol-associated DNAm changes were consistent between *in vivo* alcohol-exposed placentas and *in vitro* exposed hESCs. Prominent alcohol-induced hypomethylation was observed in most genomic locations relative to gene and CpG island (Fig. 5b) and the global DNAm level predicted by Alu, LINE1, and LTR repetitive element regions was significantly higher in LTRs of alcohol-exposed hESCs (*P* = 0.02, Student’s *t*-test) (Fig. 5c). Also, according to the GO:BP enrichment analysis, DMPs and DMRs of alcohol-exposed hESCs cluster to the GO terms involved in the function of heart and nervous system, consistently with PAE placentas (Supplementary Fig. 1 and 2, Supplementary Tables 18 and 19, respectively).

To study alcohol-induced alterations in gene expression, we performed mRNA-seq analysis for the same two hESC lines with replicates (13 alcohol-exposed and 12 control samples). A total of 4,992 genes with significantly altered expressions were observed (FDR < 0.05) (Fig. 5d and Supplementary Table 20), which are predominantly linked to the RNA processing and mitochondrial gene regulation according to the GO:BP enrichment analysis (FDR-corrected *q*-value < 0.05) (Supplementary Fig. 3 and Supplementary Table 21). The correlation analysis revealed two genes (*DPEP3, RAB17*), which had a significant PAE-associated correlation between increased DNAm and decreased mRNA expression in hESCs (Supplementary Table 22).

The expression of developmentally critical genes, pluripotency gene *SOX2* as well as both *de novo* DNA methyltransferase enzymes *DNMT3A* and *DNMT3B* (FDR = 3.52×10^−5^, FDR = 3.08×10^−11^ and FDR = 1.02×10^−8^, respectively), was significantly downregulated in alcohol-exposed hESCs (Fig. 5e). Furthermore, the ratio of *OCT4* to *SOX2* was significantly higher in alcohol-exposed hESCs compared to controls (*P* = 1.18×10^−5^, Student’s *t*-test) (Fig. 5e), which is consistent with a previous study with mouse ESCs^8^. Interestingly, it has been shown that the differentiation into the germ layers in mouse depends on the dosage of pluripotency genes *Oct4* and *Sox2*, and that *Sox2* protein level is upregulated in cells differentiating into neural ectoderm^31^.

### Effects of early *in vitro* alcohol exposure on *DPPA2* and *DPPA4* regulation

Owing to the function of *DPPA2* and *DPPA4* in the early embryonic development, we explored if alcohol exposure could affect their DNAm and expression in hESCs or after *in vitro* differentiation into the three germ layers. The genome-wide expression analysis for hESCs revealed significant alcohol-induced upregulation of *DPPA2* (FDR *=* 8.36×10^−5^), but no change in considerably actively expressed *DPPA4* (Fig. 6a). This is in line with the results of genome-wide DNAm analysis – there were two hypomethylated DMPs and one DMR in the regulatory region of *DPPA2*, but the regulatory region of *DPPA4* was unmethylated in both alcohol-exposed and control hESCs (Fig. 5a and 6b) (confirmed by EpiTYPER analysis, Supplementary Table 23). Furthermore, we studied potential alcohol-induced alterations in histone modifications H3K4me2, H3K4me3, and H3K9ac by chromatin-immunoprecipitation with quantitative PCR (ChIP-qPCR). Although the level of histone modifications in the regulatory region of *DPPA2* was very low compared to *DPPA4*, we observed increased amount of active chromatin mark H3K4me2 in the regulatory regions of both *DPPA2* and *DPPA4* as well as increased amount of H3K4me3 in *DPPA4* in alcohol-exposed hESCs (Fig. 6c). However, observed changes were not statistically significant.

**Fig. 6.**
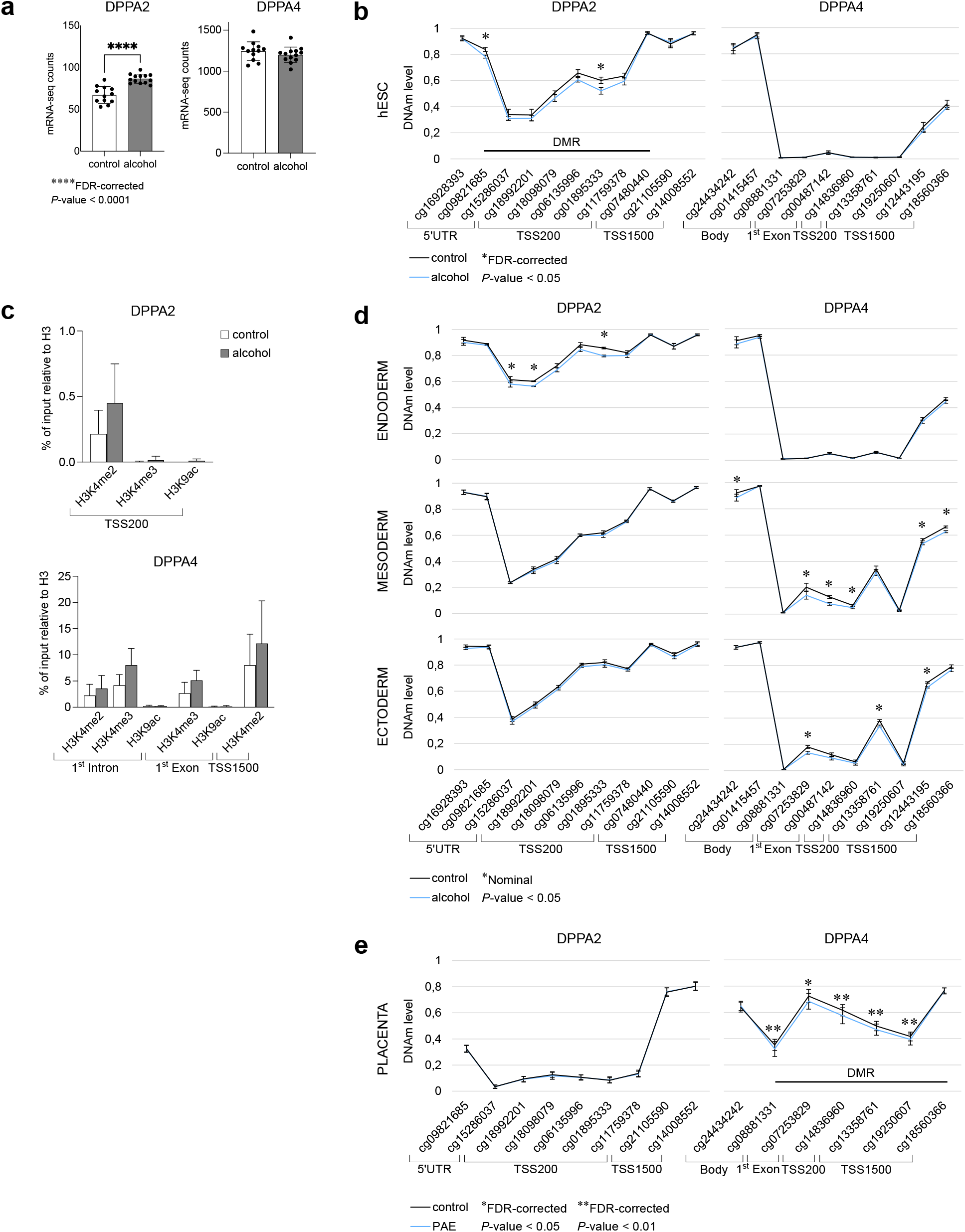
Effects of alcohol exposure on *DPPA2* and *DPPA4* in hESCs and differentiating cells. **a** *DPPA2* and *DPPA4* gene expressions in control (*n* = 12) and alcohol-exposed (*n* = 13) hESCs. **b** *DPPA2* and *DPPA4* DNAm profiles in control and alcohol-exposed hESCs (*n* = 8/condition). **c** *DPPA2* and *DPPA4* histone modifications in control and alcohol-exposed hESCs (*n* = 4/condition). Histone modification enrichments were normalized to the total histone H3. **d** *DPPA2* and *DPPA4* DNAm profiles in control and alcohol-exposed differentiated endo-, meso-, and ectodermal cells (*n* = 4/condition, respectively). **e** *DPPA2* and *DPPA4* DNAm profiles in control (*n* = 66) and PAE (*n* = 69) placentas. Data presented as mean ±SD. \**P* < 0.05, Mann-Whitney U and \*\*\*\**P* ≤ 0.0001, FDR-corrected *P*-value.

To see the potential effects of alcohol exposure on DNAm profiles of *DPPA2* and *DPPA4* regulatory regions in differentiating cells, we differentiated hESCs (H1) into the endodermal, mesodermal, and ectodermal cells *in vitro*. The cells were exposed to 70 mM alcohol during the culturing and the DNAm profiles of *DPPA2* and *DPPA4* loci were analyzed from normalized Illumina’s EPIC microarray data (four alcohol-exposed and four control samples/germ layer). The DNAm profile of endodermal cells was similar to hESCs with significant locus-specific decreased DNAm in *DPPA2* regulatory region in alcohol-exposed cells and unmethylated *DPPA4* regulatory region in both alcohol-exposed and control cells (Fig. 6d). On the contrary to the hESCs, increased methylation level in the *DPPA4* regulatory region was observed in mesodermal and ectodermal cells. Notably, consistent with the PAE placentas, both mesodermal and ectodermal cells had significant locus-specific alcohol-induced decreased DNAm in the regulatory region of *DPPA4* (Fig. 6d,e).

### Genome-wide effects of *in vivo* and *in vitro* alcohol exposure

Finally, to see the early effects of alcohol exposure on genes in both *in vivo* exposed extraembryonic placenta and *in vitro* exposed hESCs, we compared the results of genome-wide DNAm and mRNA-seq analyses (Fig. 7a and Supplementary Table 24). Only five common genes (*TRAK1, LBH, ZNF175, PRKCH*, and *TCTEX1D1*) associated with alcohol exposure in all genome-wide analyses of placenta and hESCs (differentially methylated CpGs with FDR < 0.05). A trafficking kinesin protein *TRAK1* has been associated with mitochondrial dynamics and severe neurodevelopmental disorder fatal encephalopathy^32^ as well as autism spectrum disorder (ASD)^33^, and schizophrenia^34^. *LBH regulator of WNT signaling pathway* regulates the formation of the limbs and heart in the early stage of vertebrate embryogenesis^35^. Since static encephalopathy and congenital heart defects are common features of FASD, both genes are interesting candidate genes for alcohol-induced developmental disorders.

**Fig. 7.**
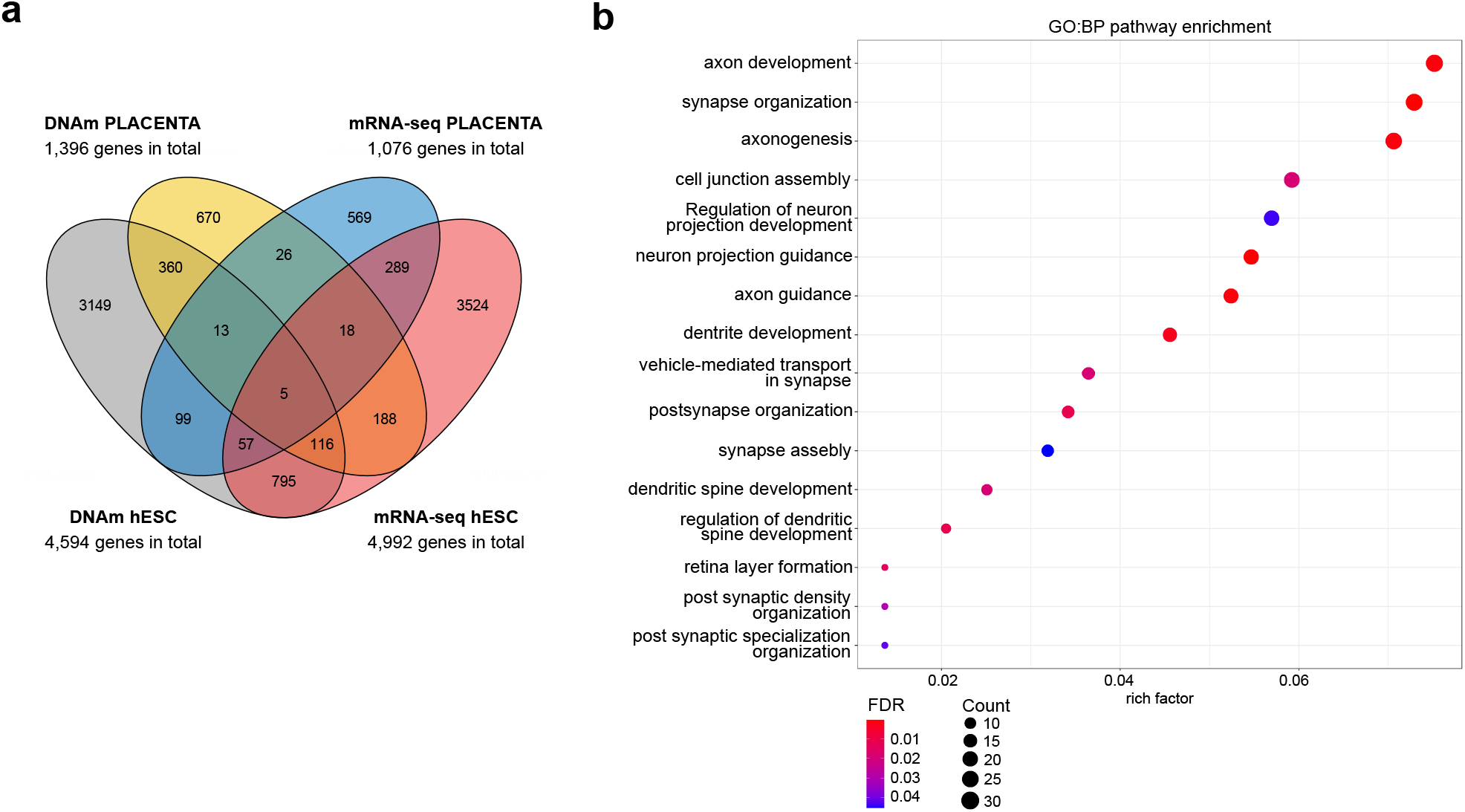
Common alcohol exposure-associated genes in the genome-wide analyses. **a** Venn diagram showing the number of common genes, which associate with alcohol exposure in the genome-wide DNAm and mRNA-seq analyses of placenta and hESCs. **b** Significantly enriched terms identified in GO:BP enrichment analysis of 494 common genes in DNAm analyses of placenta and hESCs (FDR-corrected *q*-value < 0.05). All the significant pathways are shown.

A total of 494 common genes associated significantly with alcohol exposure in DNAm analyses of placenta and hESCs. Notably, according to the GO:BP enrichment analysis, these common genes are linked exclusively to neurodevelopmental GO terms including axon development and synapse organization (FDR-corrected *q*-value < 0.05) (Fig. 7b and Supplementary Table 25). Importantly, several of these common genes have been previously associated with PAE or FASD in BECs or/and peripheral WBCs in children (Supplementary Table 26). *PTPRN2*^36^, *MAD1L1*^37^, and *AGAP1*^38^ are all linked to neurodevelopmental disorders and associate significantly with PAE or FASD in two or more previous epigenetic studies^16,17,39,40^. Furthermore, *FOXP1*, a family member of *FOXP2*, and *GLI2* have been found to associate with FASD diagnosis in BECs^40^ or WBCs^15^ in childhood, respectively. Interestingly, transcription factor *GLI2* is a mediator of Sonic hedgehog signaling, and it has been earlier associated with facial dysmorphology and brain deficiency in alcohol-exposed mouse fetuses^41^.

### Associations between candidate genes, alcohol consumption, and newborns’ phenotypes

Potential correlations between placental DNAm and gene expression of the candidate genes and maternal alcohol consumption were calculated. DMPs with the largest DNAm difference between all PAE and control placentas were examined. A moderate negative correlation between *TACR3* DNAm and alcohol units was observed in the all PAE group (cg16461251: *r* = -0,403, *P* = 0.041 and cg18538958: *r* = -0.395, *P* = 0.046, *n* = 26, Spearman’s rank correlation) and a strong negative correlation in the early PAE subgroup (cg18538958: *r* = -0.762, *P* = 0.028, *n* = 8, Spearman’s rank correlation). Furthermore, moderate correlations between AUDIT scores and *TRAK1* as well as *FOXP1* expressions (*r* = 0.384, *P* = 0.028 and *r* = 0.400, *P* = 0.021 *n* = 33, Spearman’s rank correlation, respectively) were detected in the all PAE group.

When the correlations between placental DNAm and newborns’ birth measures (SDs) and placental weight were examined, no correlations in the all PAE group were detected. However, in the early PAE subgroup, *DPPA4, FOXP2*, and *LBH* DNAm correlated moderately with the birth weight (cg14836960: *r* = -0,421, *P* = 0.026, cg18546840: *r* = -0.405, *P* = 0.033, cg07786607: *r* = 0.400 *P* = 0.035, *n* = 28, Pearson correlation, respectively). Also, *FOXP2* DNAm correlated moderately with birth weight and *LBH* DNAm with HC and placental weight (*r* = -0.452, *P* = 0.016, *r* = 0.456 *P* = 0.015, *n* = 28, Pearson correlation, *r* = 0.415, *P* = 0.028, *n* = 28, Spearman’s rank correlation, respectively).

Interestingly, the *DPPA4* DMP (cg14836960) correlated significantly with *FOXP2* (cg18546840: *r* = 0.502, *P* = 0.006, cg24786986: *r* = 0.482, *P* = 0.009, Pearson correlation, respectively) and *TACR3* (cg18538958: *r* = 0.399, *P* = 0.035, Spearman’s rank correlation, *r* = 0.429, *P* = 0.023, Pearson correlation) DMPs in the early PAE subgroup, but no correlations were observed in the all PAE placentas group. This suggests a parallel effect of early alcohol exposure on the DNAm of these three candidate genes in each placenta, which may vanish during prolonged exposure.

## DISCUSSION

Our study, which is the first genome-wide DNAm analysis of severely alcohol-exposed placentas as far as we are aware, strengthen the value of placental tissue in studying the effects of prenatal environment on human development. This can be seen as similar locus specific DNAm alterations in *DPPA4* and *TACR3* in alcohol-exposed extraembryonic and embryonic cells as well as in similar global DNAm changes in both *in vivo* and *in vitro* exposed cell types. Also, the same genes with alcohol-associated DNAm changes in placenta and hESCs were linked to the neurodevelopmental pathways. Alterations in genes involved in axon development or synapse organization may not be essential for placental cells, but DNAm changes can reveal the effects of early environment on epigenome in general, without cell or tissue specificity.

The role of *DPPA2* and *DPPA4* as chromatin remodelers and epigenetic priming factors in early development makes them plausible candidate genes for developmental disorders. Alcohol-induced alteration in the regulation of *DPPA2* in hESCs as well as decreased DNAm in *DPPA4* regulatory region in PAE placentas and alcohol-exposed mesodermal and ectodermal cells indicates that alcohol is able to affect their regulation in the early development. Delayed downregulation of epigenetic priming factors during a critical period of development can result in subtle but wide alterations in the timing and efficiency of developmental programming. Indeed, increased levels of both *DPPA2* and *DPPA4* have enhanced reprogramming to pluripotency in mouse and human cells^11^ and overexpression of *DPPA4* has been associated with inhibition of ESC differentiation into a primitive ectoderm lineage in mouse^42^. Furthermore, our study shows that alcohol exposure alters the balance of *OCT4* and *SOX2* expression in hESCs, which is in line with a previous mouse study^8^. Owing to the effects of these two lineage specifier proteins on the differentiation into mesoendoderm or neuronal ectoderm^31,43,44^, our results are consistent with the idea that alcohol can affect the cell fate decision and consequently reduce the number of ectodermal cells^8^. Observed changes in *DPPA4, OCT4*, and *SOX2* could explain the specific vulnerability of the developing nervous system to the effects of alcohol.

Another candidate gene for alcohol-induced developmental disorders identified in the current study is the transcription factor *FOXP2*, which represses genes involved in maintaining a non-neuronal state and activates genes that promote neuronal maturation by affecting chromatin structure^45^. Neuronal phenotypes associated with *FOXP2* mutations include expressive and receptive language impairment, orofacial dyspraxia, abnormalities in cortex and basal ganglia^21,46,47^ as well as attention deficit hyperactivity disorder (ADHD)^48^. Since receptive and expressive language disorders as well as ADHD have been considered as common comorbidities of FASD^49,50^, *FOXP2* is a plausible candidate gene for developmental disorders induced by PAE. Interestingly, both *FOXP1* and *FOXP2* as well as their targets have been associated with ASD^51,52,53^, which could explain the partial overlapping phenotypes between FASD, ASD, and ADHD observed in previous studies^49,54^.

The observed difference in *DPPA4* DNAm between PAE and control placentas is a potential biomarker for early alcohol exposure or even FASD, but the use of placental tissue in diagnostics has limitations. The PAE-associated trend of decreased DNAm in *TACR3* in both newborns’ placenta and BECs however suggests, that alterations are detectable in different tissue types, which makes buccal swabs a promising tool for diagnostics. Due to the ectodermal origin of BECs, buccal samples could be particularly useful for the diagnostics of neurodevelopmental disorders. Owing to the *TACR3*’s role in growth, reproduction and several processes in nervous system^55,56^ as well as the moderate correlation between placental *TACR3* hypomethylation and the number of alcohol units consumed by mothers, it is also an interesting candidate gene for alcohol-induced developmental disorders. Since genetic variation in *TACR3* has been associated with alcohol dependence^23^, potential genotypic effects on observed PAE-associated hypomethylation should be studied in the future.

By using repetitive elements, we predicted increased global alcohol-associated DNAm in both placenta and hESCs. This is consistent with the previous study, in which PAE throughout the pregnancy has been associated with higher placental global DNAm examined by using Alu repeats of male newborns^57^. Also, the increased DNAm at the LTR promoter of intracisternal A particle in the *agouti* locus was observed in our previous study, in which we showed for the first time that PAE can affect adult phenotype by altering the epigenotype of early mouse embryo^5^. However, majority of the alcohol-associated DMPs in the regulatory regions in the placenta and hESCs were hypomethylated. The mechanisms by which alcohol alters the DNAm are still mainly unknown. Enzymatic malfunction of DNMTs caused by oxidative stress or effects of alcohol on cells’ methionine cycle and consequently on DNAm level have been suggested in previous studies^58^. Also, according to previous studies the timing of exposure is fundamental – the effects differ between undifferentiated, differentiating and differentiated cells^7,59^.

We are aware of the limitations in this study. We have been able to focus only on gestational alcohol consumption, although the effects of parental alcohol consumption on gametes prior to fertilization can also affect embryonic development^60,61^, which is impossible to exclude from this study. Also, the effects of common concurrent factors such as smoking and antidepressants cannot be completely excluded. Furthermore, we need to consider that the amount and the timing of consumed alcohol is mainly self-reported by the mothers in a special outpatient clinic for pregnant women with substance use problems, and inaccuracy in this personal evaluation can occur^62^. Due to these limitations, *in vitro* experiments are necessary part of this study.

## CONCLUSIONS

By using the exceptional biological samples of PAE newborns as well as alcohol-exposed both hESCs and differentiated hESCs, our study shows the early effects of alcohol exposure on both embryonic and extraembryonic cells, reveals interesting new candidate genes *DPPA4, FOXP2*, and *TACR3* for FASD as well as brings forth potential biomarkers for PAE. The discovery of *DPPA4* and *FOXP2* introduces the role of chromatin remodelers in alcohol-induced developmental disorders. Inaccurate timing and efficiency of transcriptional programming due to unfavorable epigenetic environment could explain the wide spectrum of disorders in the FASD phenotype.

## METHODS

### epiFASD cohort

Women (*n* = 80) with abundant alcohol consumption were recruited to this study in a special outpatient clinic for pregnant women with substance use problems in Helsinki University Hospital, Finland during the years 2013–2020 (Table 1 and Supplementary Table 1). The timing of maternal alcohol consumption was registered using self-reported information. To avoid specific individual level data, the timing of consumption is presented in three categories according to pregnancy trimesters (Supplementary Table 1). The amount of maternal alcohol consumption was registered using self-reported information: Alcohol Use Disorders Identification Test (AUDIT) or the number of alcohol units per week (one unit is 12 g of ethyl alcohol). A 10-item screening tool AUDIT, developed by the World Health Organization, estimates alcohol consumption, drinking behavior, and alcohol-related problems^19^. Maternal alcohol consumption is presented in three categories according to AUDIT scores or alcohol units consumed per week^63,64^. However, self-reported information (not categories) about timing of drinking, AUDIT scores, and alcohol units were used in statistical analyses. The mothers who consumed alcohol up to gestational week seven at maximum were selected in the early PAE subgroup. Only samples with the most specified information about the maternal alcohol consumption were included (28 newborns) (Supplementary Table 1). 13 mothers of all PAE newborns did not smoked, and 18 mothers used antidepressants or antipsychotic medication during the pregnancy. Five mothers used gestational diabetes mellitus medication. Four mothers had thyroid diseases, two had antihypertensive medication, and one had preventive medication for herpes. One mother had FAS diagnosis. One mother was an occasional user of stimulants and one of cannabis. 15 (18.8%) of the deliveries were cesarean sections. Due to the preterm premature rupture of membranes, two newborns were preterm. One of the PAE newborns had two thumbs in one hand, and three had cleft lip. One newborn was Asian, one was Caucasian (other than Finnish), and two were of African origin. One’s mother was Caucasian (other than Finnish), and one had African origin father. Other newborns were children of Finnish, Caucasian parents.

The control samples (*n* = 100), collected during the years 2013–2015 in Helsinki University Hospital, Finland, were from newborns of healthy Finnish, Caucasian mothers who did not use alcohol or smoke during pregnancy according to their self-reported information (Supplementary Table 2). Ten (10%) of the deliveries were cesarean sections.

### Sample collection

Biological samples (placental biopsies, WBCs from umbilical cord blood, and BECs) of newborns were collected immediately after delivery. When this was not possible, placenta was stored in the fridge for a maximum of 12 hours and only DNA was extracted for further analyses. The placental biopsies (1 cm^4^) were collected from the fetal side of the placenta within a radius of 2–4 cm from the umbilical cord, rinsed in cold 1× PBS and stored in RNAlater^®^ (Thermo Fisher Scientific) at −80 °C. WBCs were extracted from umbilical cord blood as soon as possible, at latest 16 h after birth as described previously^65^. BEC samples were collected by rubbing buccal swabs (SK-3S, Isohelix or Catch-All™ Sample Collection Swab, Epicentre Biotechnologies) 20 times firmly against the inside of the newborn’s cheek and stored at −80 °C.

Birth weight (g), birth length (cm), and HC (cm) were examined using international growth standards, the Fenton Preterm Growth Chart by PediTools (http://peditools.org), in which the gestational age at birth and sex are considered when calculating the SD (*z*-score) of birth measures^18^.

### hESC and differentiation experiments

#### hESC culture and alcohol treatment

Alcohol concentration of 70 mM, which corresponds to the blood alcohol concentration of a heavy drinker^66^, was chosen according to a previous publication^67^. hESC lines H1 (WA01) and Regea08/017 were cultured in E8 or in E8 Flex Medium (Gibco) on Matrigel (Corning) coated plates at 37 °C and 5% CO2. Culture media was routinely replaced every day (every second or third day in the case of E8 Flex Medium) and cells were passaged using 0.5 mM EDTA. For the alcohol treatment, the medium was supplemented with alcohol (≥ 99.5 p-% ethanol) at a final concentration of 70 mM 48 h before the cells reached confluency and were cross-linked for ChIP or collected for DNA and RNA extractions. Due to alcohol evaporation, the culture media with alcohol were replaced after treatment of 24 h.

#### Germ layer cell differentiation and alcohol treatment

H1 cells cultured in E8 Medium on Matrigel plates were differentiated into the endodermal, mesodermal, and ectodermal cells by using the STEMdiff™ Trilineage Differentiation Kit (StemCell Technologies, Inc.). Cells were seeded on a Matrigel-coated 6-well plates at 250,000 cells/well for the mesoderm, 1 million cells/well for the endoderm and 1.5 million cells/well for the ectoderm, and differentiated according to manufacturer’s instructions. The cells were supplemented with 10 µM Y-27632 for the first 24 h after seeding, and the mediums were changed daily. For the alcohol-exposed wells, the medium was supplemented with alcohol at a final concentration of 70 mM. After 5 or 7 days the cells were collected for DNA and RNA extractions. The differentiation was confirmed by mRNA-seq and expression profiles of genes characteristic for specific germ layers were analyzed (Supplementary Fig. 4).

### DNA and RNA extractions

Placental genomic DNA was extracted from one to four (3.7 on average) pieces of placental tissue samples using commercial QIAamp Fast DNA Tissue Kit (Qiagen) or standard phenol-chloroform protocol. WBC DNA was extracted using QIAamp Fast DNA Tissue Kit or AllPrep DNA/RNA/miRNA Universal Kit (Qiagen) and BEC DNA using BuccalPrep Plus DNA Isolation Kit (Isohelix). Placental RNA was extracted from the same pieces as DNA (2.9 on average) by AllPrep DNA/RNA/miRNA Universal Kit, and the same kit was used for DNA and RNA extraction from hESCs and differentiated hESCs. RNA quality was assessed using an Agilent 2100 Bioanalyzer (Agilent Technologies, Inc.), which was provided by the Biomedicum Functional Genomics Unit (FuGU) at the Helsinki Institute of Life Science and Biocenter Finland at the University of Helsinki.

### DNAm microarrays

Genomic DNA (1,000 ng) from available placental (all PAE *n* = 69, early PAE *n* = 28, and control *n* = 66), hESC (H1 and Regea08/017: *n* = 4/condition, respectively), and differentiated hESC (each germ layer: *n* = 4/condition) samples was sodium bisulfite-converted using the Zymo EZ DNAm™ kit (Zymo Research) and genome-wide DNAm was assessed with Infinium Methylation EPIC BeadChip Kit (Illumina) following a standard protocol.

#### DMP analysis

The raw DNAm dataset was pre-processed using ChAMP R package^68^. The data filtering steps included the removal of probes located in sex chromosomes and probes binding to polymorphic and off-target sites^69^. For placental samples, probes located in Finnish specific SNPs (SNPs which overlap with any known SNPs with global minor allele frequency (MAF) and MAF in a Finnish population > 1%) were removed as described previously^70^. Population specific masking and SNP information was obtained from Zhou et al.^71^. Type-I and Type-II probes were normalized using the BMIQ method implemented in ChAMP. Potential batch effects caused by technical factors and biological covariates were studied from singular value decomposition plots. For placental samples, the batch-effect correction was performed by the Empirical Bayes method using the R package ComBat^72^ and the model was adjusted to take into account for biological covariates sex and smoking. For hESCs, a mixed linear model was built by using humanzee R package (http://giladlab.uchicago.edu) to remove sample-specific random effect. Subsequently, a total of 588,781 probes of placental and 800,002 probes of hESC samples were retained for further downstream analysis. DMP analysis by using M-values was performed by R package Limma^73^ and the model for placental samples was adjusted to consider biological covariates sex and smoking. *β*-values were used for visualization and interpretation of the results and to construct the DNAm profiles of differentiated cells. DMPs were considered as significant when DNAm difference was greater than 5% (−0.05 ≥ Δβ ≥ 0.05) and FDR-corrected *P*-value smaller than 0.05. Benjamini-Hochberg procedure was used to control for FDR.

#### DMR analysis

DMRcate R package was used for analyzing DMRs^74^. The method uses minimum description length for detecting region boundaries in DMR identification. DMRcate was adjusted to determine probes (≥ 3) in a region with maximal allowed genomic distance of 1,000 bp containing at least one CpG with Δβ ≥ 5%. Further, FDR < 0.05 was defined to describe the DMR with significance.

#### Pathway analysis

Enrichment analysis was performed for significant DMPs by *gometh* function in missMethyl R package^75^, which considers the different number of probes per gene present on the EPIC array and CpGs that are annotated to multiple genes. missMethyl was set to use the GO knowledgebase as the source for identifying significantly enriched BP terms from genes which contained at least one significant DMP. Pathway analysis was also performed for significant DMRs by *goregion* function in missMethyl R package and GO:BP knowledgebase was used as a source. For the enrichment analysis, DMRs with two CpG sites were also included.

#### Global methylation

Filtered and corrected DNAm data was used to predict DNAm in Alu, LINE1, and LTR using Random Forest-based algorithm implemented by REMP R package^25^ as a proxy for global DNAm level. Less reliable predicted results were trimmed according to quality score threshold 1.7 and missing rate 0.2 (20%).

### EpiTYPER

To validate and replicate the findings from the EPIC microarrays, DNAm profiles of target genes (*DPPA4, FOXP2*, and *TACR3*) were measured by MassARRAY EpiTYPER (Agena Bioscience, Inc.) in placental tissue, WBCs, BECs, and hESCs. Samples of 16 PAE and 14 control newborns were chosen for the *DPPA4* target gene and 10 PAE and 9 control newborns for other target genes. In hESCs, two biological replicates of alcohol-exposed and control cells of both H1 and Regea08/017 cell lines were used for the analysis. First, genomic DNA (500–1,000 ng) was subjected to sodium bisulfite conversion using EZ-96 DNA Methylation™ kit (Zymo Research). PCR amplification was performed in three independent 10 or 15 µl reactions using HotStar PCR kit (Qiagen) following the provider’s instructions. Primers for the target regions were designed using EpiDesigner software (Agena Bioscience, Inc.: http://www.epidesigner.com) incorporating CpGs chosen for each target according to the microarray analysis. Primers for *TACR3* DMP cg18538958 with the largest DNAm difference between PAE and control groups were unable to design and therefore a correlating probe cg16461251 (*r* = 0.973, *P* < 0.001, *n* = 136, Spearman’s rank correlation) was selected for the analysis. Primers and PCR protocols for each target sequence are presented in Supplementary Tables 27 and 28. Owing to the proximity of two CpGs in one unit in *FOXP2*, they were analyzed together as the mean DNAm value. Technical replicates showing > 5% from the median value were excluded and the DNAm values from the remaining two replicates were used in the further analyses. Samples with two or three unsuccessful replicates were excluded.

### 3’mRNA-seq analysis

#### Differential expression analysis

Drop-seq pipeline^76^ was used to construct the mRNA-seq count table for available placental (all PAE *n* = 64, early PAE *n* = 23, and control *n* = 41), hESC (H1: alcohol-exposed *n* = 7 and control *n* = 6, Regea08/017: alcohol-exposed and control *n* = 6, respectively), and differentiated hESC (control endoderm *n* = 4, mesoderm *n* = 3, and ectoderm *n* = 4) RNA samples provided by FuGU. A total of 38,434 genes were identified for downstream analysis of placental and 30,081 genes of hESC samples. Principal component analysis implemented in DESeq2^77^ was used to identify batch effects and ComBat-seq^78^ to adjust separate mRNA-seq batches. Differential expression analysis was performed by DESeq2 R package, with model adjusting for smoking and sex for placental samples and with model adjusting cell line for hESCs. Same differential expression analysis method for placental samples was also used without covariates. Genes were considered as differentially expressed when FDR-corrected *P*-value was < 0.05. Benjamini-Hochberg procedure was used to control for FDR. To validate the hESC differentiation into the germ layer cells, normalized counts of marker genes were used in heatmap visualization.

#### Pathway analyses

*enrichgo* function in R package clusterProfiler version 4.0^79^ was used to perform gene-set enrichment analysis for significant differentially expressed genes. The GO knowledgebase was used as the source for identifying significantly enriched BP terms (FDR-corrected *q*-value < 0.05). Benjamini-Hochberg procedure was used to control for FDR.

### Correlation analysis

Normalized genome-wide DNAm data was compared to similarly adjusted mRNA-seq data to discover genes, which DNAm changes correlate with mRNA expression in the placenta and hESCs. A total of 53 PAE and 39 control placental samples as well as eight alcohol-exposed and eight control hESC samples, of which DNAm and mRNA expression data were available, was used. DNAm and expression data were adjusted to include only the same identified genes between the analyses. For placental data, a total of 126,810 probes were clustered according to 14,635 genes, which were identified from the mRNA-seq data. For hESCs, 106,341 probes were clustered according to 14,051 genes. MethylMix version 2.20.0. R package^80^ was used to perform correlation analysis.

### Common genes in genome-wide DNAm and mRNA-seq analyses

The gene name annotation information from DNAm (all differentially methylated CpGs with FDR < 0.05) and mRNAseq (FDR < 0.05) analyses of placenta and hESCs were used to explore the common genes that associate significantly with alcohol exposure. When CpG was annotated to multiple genes, the first UCSC gene name were chosen. If the UCSC gene name was missing, the GENCODE database information was used. GO:BP enrichment analysis of the common genes was performed by R package clusterProfiler version 4.0 (see 3’mRNA-seq pathway analysis).

### ChIP-qPCR

ChIP was performed for ∼5 million hESCs as described in Schmidt et al.^81^ with some modifications using H1 and Regea08/017 cell lines. Two replicates of both cell lines were used, which is four replicates of alcohol-exposed and control ChIP samples in total. Briefly, cells were cross-linked using 1% formaldehyde and sonicated with Bioruptor^®^ Pico sonication device (Diagenode) using optimized parameters 4 cycles of 30s on/90s off to generate DNA fragments of 300–600 bp. For immunoprecipitation, 0.75 mg of Dynabeads™ Protein G magnetic beads (Invitrogen) were first incubated with 5 µg of antibodies against H3K4me3, H3K4me2, H3K9ac, and H3 (Supplementary Table 29). Subsequently, the shared chromatin was incubated with antibody-bound protein G beads overnight at 4 °C with rotation. The protein-DNA complexes were then washed, eluted, reverse cross-linked, and treated with Proteinase K and RNase A (Thermo Scientific). Finally, the DNA was purified using QIAquick PCR Purification Kit (Qiagen) and used as a template for qPCR. The qPCR was performed in triplicates of 10 µl reactions using SsoAdvanced™ Universal SYBR^®^ Green Supermix (Bio-Rad Laboratories) according to the manufacturer’s instructions. The enrichment was normalized against input and further against total histone H3 enrichment. To compare the enrichment between alcohol-exposed and control samples, the data were also normalized against a negative control region designated as ‘Gene desert’. Target sequences were designed to incorporate regions at *DPPA2* and *DPPA4* enriched with histone modifications of interest in the H1 hESC line according to Encyclopedia of DNA Elements (ENCODE) (https://encodeproject.org/). Primers for the target sequences were designed using Primer3^82^ and primers for the negative control region were obtained from a previous publication^83^. Primers and location of amplicons in the genome, as well as qPCR protocol, are provided in Supplementary Tables 27 and 28.

### Statistical analysis

All statistical analyses were conducted using IBM SPSS Statistics for Windows, version 27.0 (IBM Corp.) or GraphPad Prism 9 software (GraphPad Software, Inc.). All data are expressed as the mean with ±SD for a normal distribution of variables. Statistical analyses were performed as described in the figure legends or in the relevant sections.

## Data Availability

The datasets supporting the conclusions of the current study are included within the article and its additional files. Due to the sensitive nature of the patient data used, the data sets are not and cannot be made publicly available. All data produced in the present study are available upon reasonable request to the authors.

## AVAILABILITY OF DATA AND MATERIALS

The datasets supporting the conclusions of the current study are included within the article and its additional files. Due to the sensitive nature of the patient data used, the data sets are not and cannot be made publicly available.

## ACKNOWLEDGEMENTS

We are grateful to all the families who took part in this study. We would also like to acknowledge research nurse Teija Karkkulainen for her contribution to this work and Dr Sailalitha Bollepalli for her bioinformatic assistance.

## CONTRIBUTIONS

P.A., J.V., T.O., H.S., M.O., T.R. and N.K-A. contributed to the study design. H.K. recruited the study participants. P.A., J.V., H.M., E.W., L.A., and N.K-A. contributed to the sample collection and processing. P.A., J.S., E.W. and N.K-A. contributed to the laboratory experiments. P.A., J.V., H.M., M.V., A.S., M.O., and N.K-A. contributed to the data analysis. P.A. and N.K-A. drafted the manuscript. All authors contributed to the revision of the manuscript. All authors gave final approval of the version to be published.

## ETHICS DECLARATIONS

### Ethics approval and consent to participate

Informed consent was obtained from all participants and the study was approved by the Ethics Committee of Helsinki University Central Hospital (386/13/03/03/2012). hESC cell line H1 (WA01) was obtained from Biomedicum Stem Cell Center (BSCC, Helsinki, Finland) through a license agreement with WiCell, Inc. and Regea08/017 from Skottman laboratory (Tampere, Finland) approved by the Ethics Committee of Tampere University Hospital (Skottman/R05116).

## SUPPLEMENTARY MATERIAL

### Supplementary Information

Supplementary Figures 1–4

### Supplementary Tables

Supplementary Tables 1–29

