## Supplementary Information for "Chromatin remodeler *developmental pluripotency associated factor 4* (*DPPA4*) is a candidate gene for alcohol-induced developmental disorders"

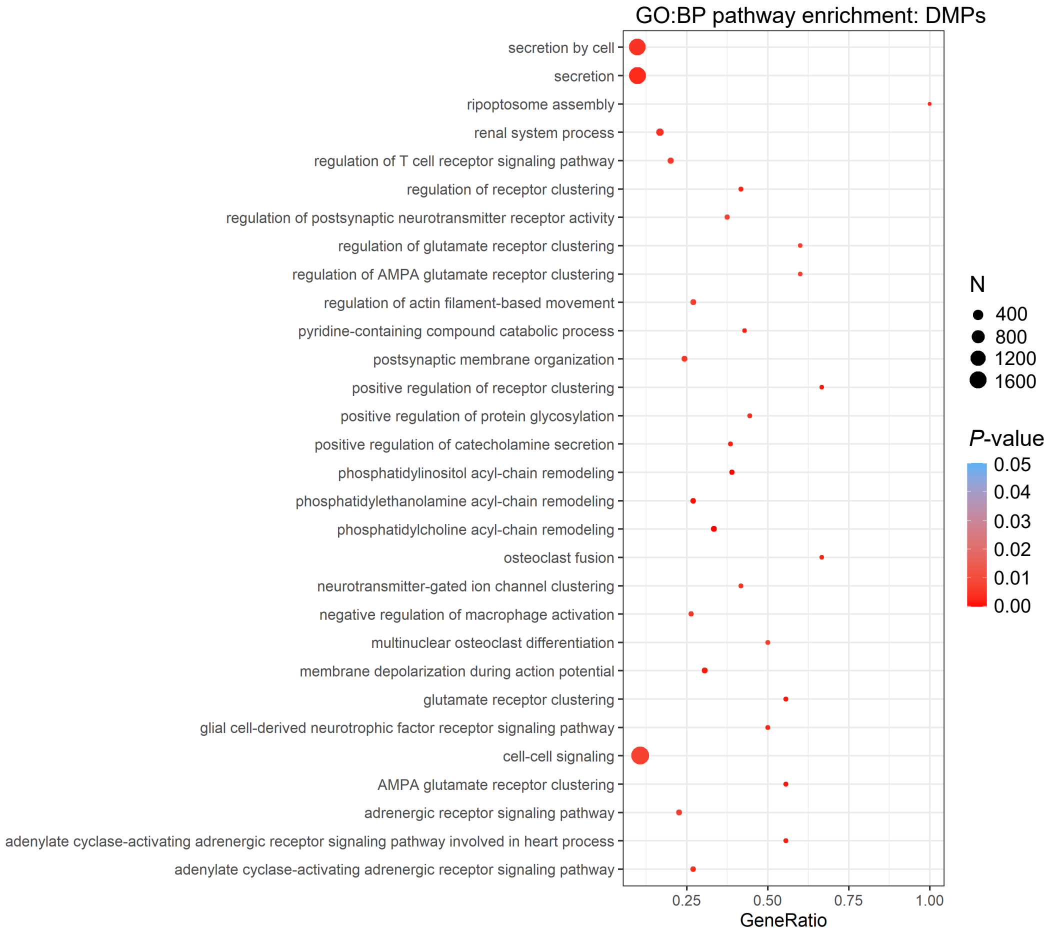


**Supplementary Figure 1: Pathway analysis of DMPs in hESCs.** Significantly enriched terms identified in GO:BP enrichment analysis of alcohol-induced DMPs in hESCs (*P* < 0.05). The 30 most significant pathways are shown.

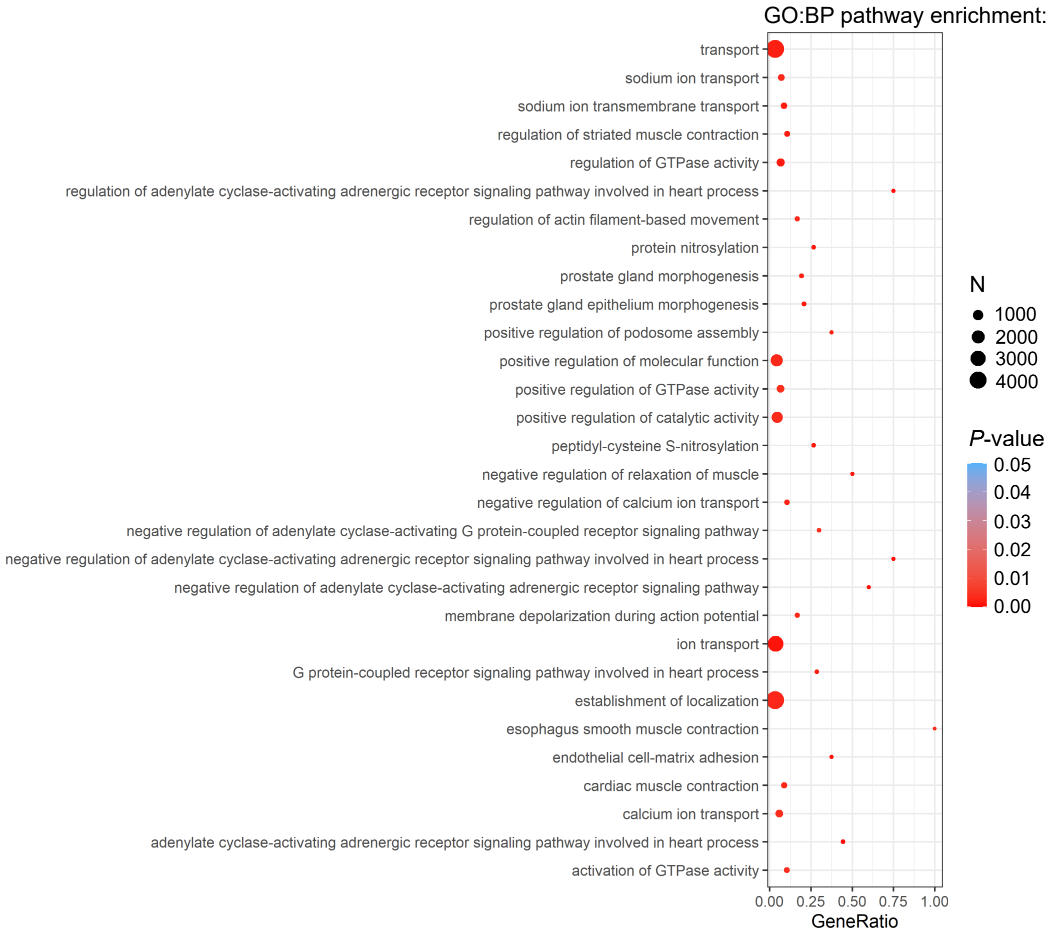


**Supplementary Figure 2: Pathway analysis of DMRs in hESCs.** Significantly enriched terms identified in GO:BP enrichment analysis of alcohol-induced DMRs in hESCs ($P < 0.05$). The 30 most significant pathways are shown.

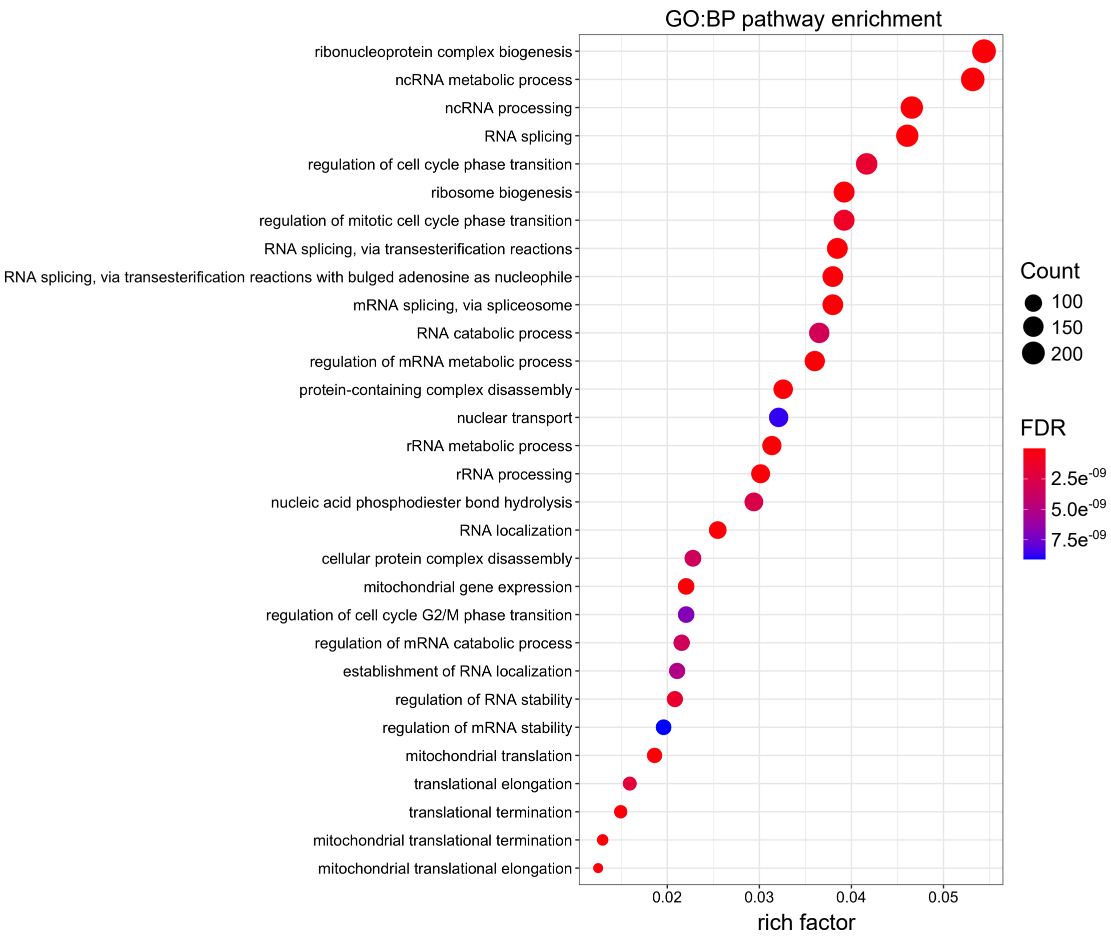


**Supplementary Figure 3: Pathway analysis of differentially expressed genes in hESCs.** Significantly enriched terms identified in GO:BP enrichment analysis of alcohol-induced differentially expressed genes in hESCs (FDR-corrected $q$-value < 0.05). The 30 most significant pathways are shown.

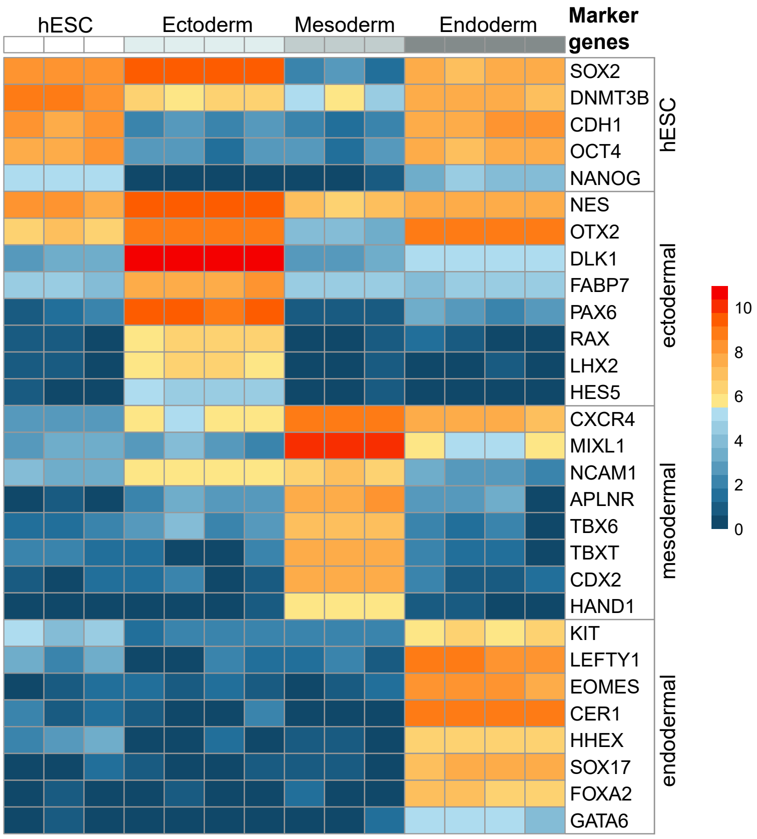


**Supplementary Figure 4: Characterization of hESC differentiation into germ layer cells.** Heatmap visualization of cell type specific gene expression profiles in undifferentiated hESCs (*n* = 3) and differentiated ectodermal (*n* = 4), mesodermal (*n* = 3), and endodermal (*n* = 4) cells.
